# Baseline ESR–CRP difference (D score) and JAK inhibitor discontinuation for loss of efficacy after biologic failure in rheumatoid arthritis

**DOI:** 10.64898/2026.01.10.26343860

**Authors:** Daisuke Oryoji, Goro Doi, Sho Fujimoto, Naoya Nishimura, Ayako Kuwahara, Masahiro Ayano, Yasutaka Kimoto, Hiroaki Niiro, Hiroki Mitoma

**Author notes:** Correspondence to: Hiroki Mitoma, MD, PhD, Department: Department of Internal Medicine, Institution: Kyushu University Beppu Hospital, Street address: 4546 Tsurumibara, City: Beppu, Postal code: 874-0838.

## Abstract

**Objectives:** To assess whether baseline ESR–CRP difference (D score) is associated with discontinuation due to loss of efficacy during Janus kinase inhibitor therapy after biologic failure in rheumatoid arthritis.

**Methods:** Single-centre retrospective cohort of 24 patients with rheumatoid arthritis who initiated a Janus kinase inhibitor after inadequate response to at least one biologic DMARD. D score was ESR (mm/h) minus CRP (mg/L). The primary outcome was discontinuation due to loss of efficacy; other discontinuations were censored. Kaplan–Meier curves and Cox models were used.

**Results:** Seven patients discontinued due to loss of efficacy. After dichotomisation at the median D score (20.3; n = 12 per group), 1-year LOE-free persistence was 90.9% in the high D group and 43.2% in the low D group (log-rank p = 0.004). The hazard ratio per 10-unit increase was 0.47 (95% CI 0.29 to 0.76; p = 0.002) and 0.41 after age adjustment (0.22 to 0.74; p = 0.003).

**Conclusions:** Baseline D score was associated with lower risk of discontinuation due to loss of efficacy. Larger studies are needed.

**Key messages:**

- Loss-of-efficacy discontinuation tended to cluster in patients with low ESR and low CRP at baseline.
- Higher baseline D score was associated with fewer loss-of-efficacy discontinuations during JAK inhibitor therapy.
- D score from ESR and CRP may complement post-biologic treatment decisions, pending external validation.

## Introduction

Rheumatoid arthritis (RA) is a chronic autoimmune inflammatory disease that can lead to irreversible joint damage and disability. The treat-to-target strategy and the introduction of biologic and targeted synthetic disease-modifying antirheumatic drugs (bDMARDs and tsDMARDs) have improved outcomes, yet a subset of patients experience persistent inflammatory activity, disabling symptoms, or repeated treatment failure. The European Alliance of Associations for Rheumatology (EULAR) has proposed a definition for difficult-to-treat RA (D2T RA) that captures patients with prior failure of multiple b/tsDMARDs and ongoing disease that remains problematic for the patient and clinician.[1–3] In routine care, patients with prior biologic failure often face limited remaining options and substantial unmet need.

Janus kinase inhibitors (JAKi), a class of tsDMARDs, are established options after inadequate response to bDMARDs, supported by EULAR recommendations and randomised trials in biologic-refractory populations.[4–7] However, real-world treatment persistence varies, and reasons for discontinuation frequently include loss of efficacy (LOE) rather than toxicity alone. Reliable predictors of sustained benefit are lacking, so therapeutic sequencing after biologic failure is commonly driven by prior treatment history, comorbidities, safety considerations, and patient preferences. Pragmatic biomarkers that are routinely available at treatment initiation could help identify patients at higher risk of early LOE and could inform counselling and follow-up intensity.

RA exhibits heterogeneity in inflammatory pathways. Type I interferon (IFN) activity and interferon-regulated gene signatures have been associated with autoantibody production, disease phenotypes, and variable responses to biologic therapies. Because JAK inhibition can modulate interferon-related signalling, an IFN-high phenotype has been hypothesised to respond differentially to JAKi in some settings. Direct assessment of IFN activity, however, is rarely feasible in routine practice.[8–14] We therefore focused on laboratory surrogates derived from standard acute-phase markers. Erythrocyte sedimentation rate (ESR) and C-reactive protein (CRP) reflect overlapping but partly distinct biology: ESR is influenced by immunoglobulins, fibrinogen, and anaemia, whereas CRP more directly reflects hepatic acute-phase responses that are often mediated by interleukin-6. ESR and CRP are not always concordant within individuals, suggesting that their combined pattern may convey information beyond either marker alone.

We operationalised the balance between ESR and CRP using a simple D score defined as ESR (mm/h) minus CRP (mg/L), selected for ease of calculation in routine care. In this single-centre retrospective cohort of biologic-experienced patients with RA initiating JAKi after inadequate response to at least one bDMARD, we evaluated whether baseline D score was associated with discontinuation due to LOE and compared its discriminative performance with ESR and CRP.

## Methods

We conducted a single-centre retrospective cohort study at Kyushu University Beppu Hospital. Consecutive adults with rheumatoid arthritis meeting the 2010 ACR/EULAR criteria who started a Janus kinase inhibitor after inadequate response to at least one biologic DMARD between April 2018 and 20 November 2025 were included. Baseline was the date of Janus kinase inhibitor initiation. Erythrocyte sedimentation rate (ESR; mm/h) and C-reactive protein (CRP; mg/dL) measured on that date or the closest values within 14 days before initiation were extracted. CRP was converted to mg/L by multiplying by 10. We prespecified a D score as ESR (mm/h) minus CRP (mg/L), a pragmatic laboratory index capturing the balance between ESR and CRP. All included patients had baseline ESR and CRP available; no imputation was performed.

The primary outcome was discontinuation due to loss of efficacy (LOE), defined as treatment discontinuation documented by the treating rheumatologist due to insufficient clinical response. Standardised disease activity scores were not consistently available in routine practice and were therefore not used to adjudicate LOE. Discontinuations for reasons other than LOE, including adverse events, were censored at the stop date (cause-specific approach). Ongoing treatments were censored at the data cutoff date (20 November 2025). Time to LOE-related discontinuation was evaluated using Kaplan–Meier curves and compared using the log-rank test. For visualisation, patients were dichotomised at the cohort median D score. Cox proportional hazards models were fitted with biomarkers scaled per 10 units, and proportional hazards assumptions were assessed using Schoenfeld residuals. In exploratory analyses, discrimination was evaluated using Harrell’s C-index and 1-year AUC, with 95% confidence intervals obtained by non-parametric bootstrap resampling (1000 iterations); further details are provided in the Supplementary Methods. Statistical analyses were performed using Python 3 with pandas, NumPy, SciPy, lifelines and scikit-learn. Figures were generated using Matplotlib. The institutional ethics committee approved the study with waiver of informed consent because of the retrospective design and use of anonymised data. Additional analyses are provided in Supplementary Tables S1 to S8 and Supplementary Figures S1 to S5.

## Results

### Patient characteristics at JAK inhibitor initiation

Twenty-four patients with rheumatoid arthritis who initiated a Janus kinase inhibitor after inadequate response to at least one prior biologic DMARD were included. The median age at initiation was 71.0 years (interquartile range [IQR] 57.8 to 75.0), and 21 patients (87.5%) were female. Index Janus kinase inhibitors were baricitinib (n = 12), upadacitinib (n = 7), and filgotinib (n = 5). The median baseline erythrocyte sedimentation rate (ESR) was 34.0 mm/h (IQR 10.0 to 69.8), the median C-reactive protein (CRP) was 4.3 mg/L (IQR 0.3 to 22.4), and the median D score was 20.3 (IQR 3.4 to 43.0). Median follow-up was 358 days (IQR 183 to 754). During follow-up, seven patients (29.2%) discontinued treatment due to loss of efficacy and two patients (8.3%) discontinued due to adverse events; adverse-event discontinuations were treated as censoring events at the stop date in the primary analysis. Baseline characteristics are shown in Table 1, and baseline characteristics stratified by D score group are shown in Supplementary Table S1.

**Table 1.**
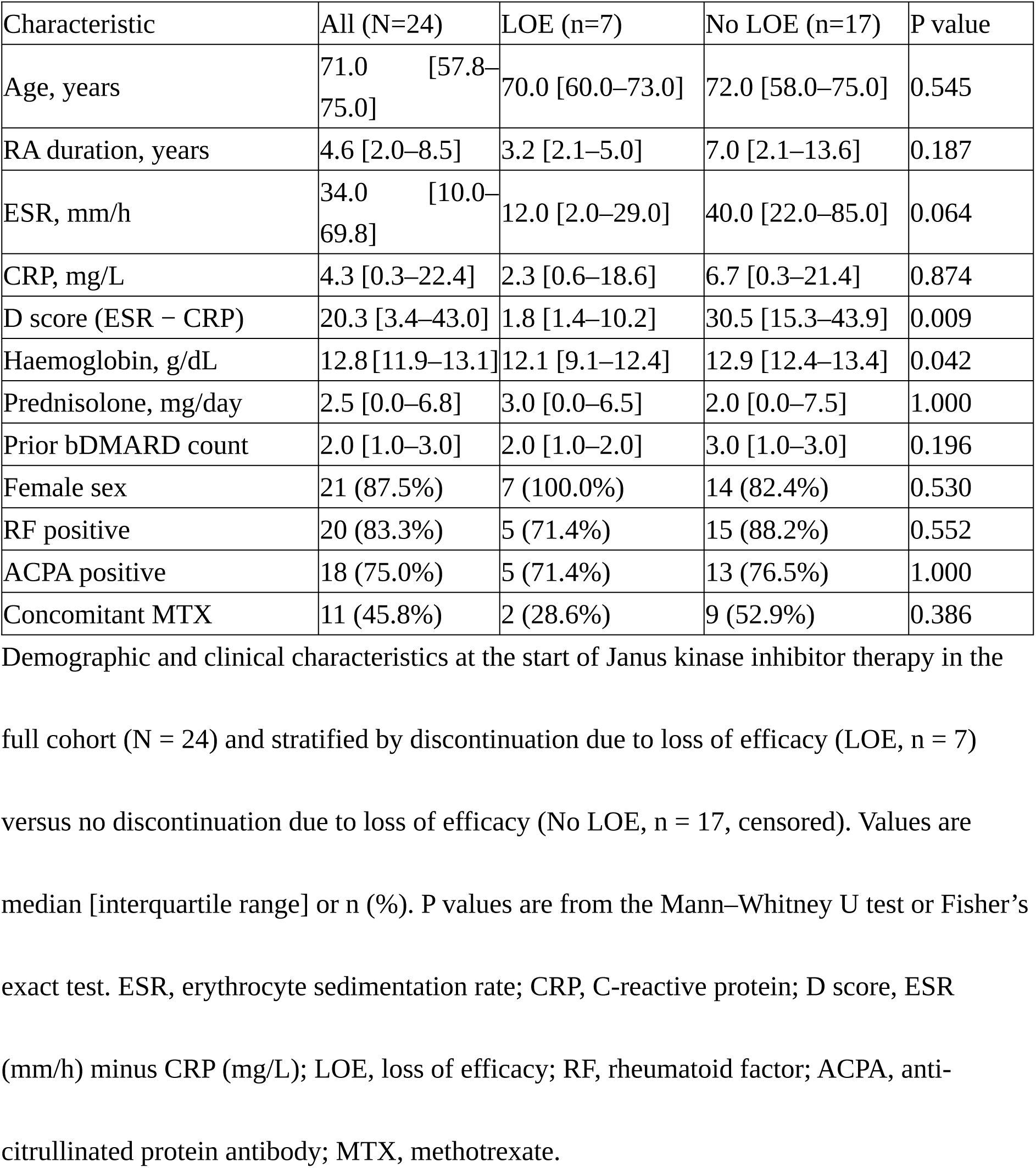
Baseline characteristics of patients at initiation of JAK inhibitor therapy.

### Association between D score and risk of discontinuation

In univariable Cox models, a higher baseline D score was inversely associated with discontinuation due to LOE (hazard ratio 0.47 per 10-unit increase; 95% confidence interval 0.29 to 0.76; p = 0.002). ESR showed a weaker association (hazard ratio 0.73 per 10 mm/h; 95% confidence interval 0.52 to 1.02; p = 0.066), whereas CRP showed no association (hazard ratio 1.00 per 10 mg/L; 95% confidence interval 0.73 to 1.36; p = 0.984). After adjustment for age, the inverse association between D score and discontinuation remained (hazard ratio 0.41; 95% confidence interval 0.22 to 0.74; p = 0.003) (Supplementary Tables S2 and S3).

### LOE-free persistence according to D score group

For visualisation, patients were dichotomised at the cohort median D score of 20.3 (high D group, n = 12; low D group, n = 12). LOE-free persistence was longer in the high D group than in the low D group. One-year LOE-free persistence was 90.9% in the high D group versus 43.2% in the low D group (log-rank p = 0.004) (Figure 1).

**Figure 1.**
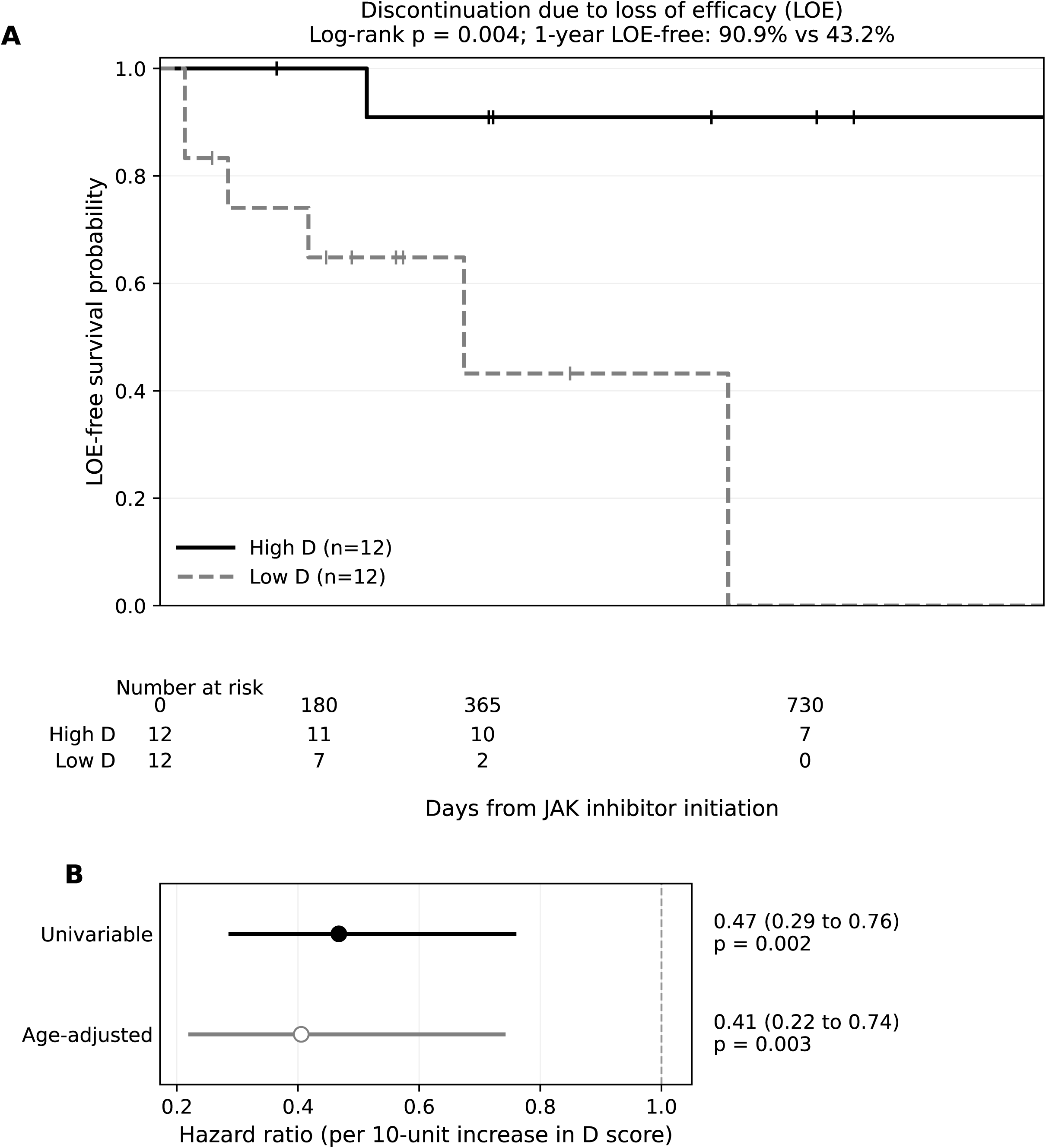
LOE-free persistence and hazard ratios by baseline D score. (A) Kaplan–Meier curves for discontinuation due to loss of efficacy, stratified by median D score (high D: n = 12; low D: n = 12; log-rank p = 0.004). Tick marks denote censored observations. Numbers at risk are shown below. (B) Hazard ratios from Cox models for loss-of-efficacy discontinuation; values below 1 indicate lower risk. D score was calculated as ESR (mm/h) minus CRP (mg/L).

### Discriminative performance of inflammatory markers

In exploratory analyses, D score showed numerically higher discrimination than ESR or CRP (Supplementary Table S4; Supplementary Figure S1).

Additional and sensitivity analyses are provided in the Supplementary Data. Proportional hazards diagnostics showed no major departures, but power was limited by the small number of events (Supplementary Figure S2). An all-cause discontinuation analysis (LOE or adverse events) yielded qualitatively similar separation between D score groups (Supplementary Figure S3), and adjustment for haemoglobin did not materially change the association (Supplementary Table S5). In an exploratory ESR×CRP quadrant analysis, discordant patterns were uncommon and LOE discontinuations tended to occur more often among patients with concordantly low ESR and CRP (Supplementary Table S6; Supplementary Figures S4 and S5). Sensitivity analyses considering preceding bDMARD classes and exposure to an IL-6 receptor inhibitor are shown in Supplementary Tables S7 and S8.

## Discussion

In this small exploratory retrospective cohort of biologic-experienced patients with rheumatoid arthritis who started a Janus kinase inhibitor after inadequate response to at least one biologic DMARD, baseline ESR–CRP difference (D score) was associated with a lower risk of discontinuation due to loss of efficacy. In exploratory analyses, D score showed numerically higher discrimination than ESR or C-reactive protein (Supplementary Table S4).

Although we did not measure type I interferon (IFN) activity, ESR and CRP reflect partly distinct inflammatory biology. ESR is influenced by immunoglobulins, fibrinogen and anaemia, whereas CRP predominantly reflects interleukin-6 driven hepatic synthesis. Because JAK inhibition can downregulate IFN related pathways,[8] ESR–CRP discordance may capture inflammatory features relevant to JAKi response, but this requires confirmation with direct IFN assessments.[9–14] Alternatively, discordance may reflect inflammation that is not primarily interleukin-6 driven and may be influenced by prior interleukin-6 receptor inhibition. Disentangling these mechanisms will require larger cohorts with detailed treatment histories and direct interferon measurements.

Our cohort reflects real-world decision making after biologic failure and includes patients with features overlapping difficult-to-treat RA (D2T RA).[1–3] If validated, the D score could provide an accessible biomarker to complement clinical judgment when choosing among b/tsDMARD options.

Because the endpoint focused on discontinuation due to loss of efficacy, the D score may be most relevant to anticipating insufficient response rather than discontinuation for adverse events. In practice, the score could be considered alongside comorbidities and safety considerations when selecting a Janus kinase inhibitor versus alternative b/tsDMARDs after biologic failure, but it should not be used as a sole decision tool. Strengths of this study include consecutive patient inclusion, a prespecified score definition and transparent reporting of effect estimates and discrimination metrics.

Limitations include small sample size and few events, with risk of overfitting and optimistic discrimination metrics. Because only seven events occurred, we limited multivariable adjustment to age to reduce overfitting. Residual confounding is possible, ESR and CRP are non-specific, and haemoglobin differed between outcome groups; anaemia can increase ESR, and the D score may partly capture anaemia-related ESR elevation rather than treatment-specific biology. In a sensitivity analysis including haemoglobin, the association between D score and discontinuation remained in the protective direction, although estimates were attenuated (Supplementary Table S5). In an additional sensitivity analysis using all-cause discontinuation as the event, separation between D score groups remained (Supplementary Figure S3). Longitudinal disease activity and safety outcomes were not comprehensively captured. We also lacked direct IFN measurements. Adverse event discontinuations were treated as censoring events in the primary analysis and may act as a competing risk; given the small number of adverse event discontinuations, we could not perform formal competing-risk modelling. Therefore, results pertain to discontinuation due to loss of efficacy rather than all-cause discontinuation. We could not formally adjust for individual JAK inhibitors because of the limited number of events.

Overall, ESR–CRP discordance summarised by D score may help identify biologic-experienced patients more likely to remain on JAK inhibitors. These findings should be considered exploratory and require external validation before any clinical application. Larger multicentre cohorts with interferon related biomarkers and detailed prior biologic exposure should validate these findings and clarify clinical utility.

## Acknowledgements

We thank the staff at the Department of Internal Medicine, Kyushu University Beppu Hospital, for their support in data collection. Generative AI assistance (ChatGPT, OpenAI) was used for language editing. The authors reviewed and take responsibility for the content.

## Author Contributions

DO drafted the manuscript. All authors contributed to the study design, data analysis, and critical revision of the manuscript. All authors read and approved the final manuscript.

## Data Availability Statement

The data underlying this article cannot be shared publicly due to the privacy of individuals that participated in the study. The data will be shared on reasonable request to the corresponding author.

## Ethics approval

The study was approved by the institutional review board of Kyushu University (Approval No. 25013). Informed consent was waived due to the retrospective nature of the analysis. Patient and public involvement: Patients and/or the public were not involved in the design, conduct, reporting, or dissemination plans of this research.

## References

1. Nagy G, Roodenrijs NMT, Welsing PMJ, Kedves M, Hamar A, van der Goes MC, et al. EULAR definition of difficult-to-treat rheumatoid arthritis. Ann Rheum Dis. 2021;80(1):31–35.

2. Watanabe R, Okano T, Gon T, Yoshida N, Fukumoto K, Yamada S, et al. Difficult-to-treat rheumatoid arthritis: current concept and unmet needs. Front Med (Lausanne). 2022;9:1049875.

3. Hofman ZLM, Roodenrijs NMT, Nikiphorou E, Kent AL, Nagy G, Welsing PMJ, et al. Difficult-to-treat rheumatoid arthritis: what have we learned and what do we still need to learn? Rheumatology (Oxford). 2025;64(1):65–73.

4. Smolen JS, Landewé RBM, Bergstra SA, Kerschbaumer A, Sepriano A, Aletaha D, et al. EULAR recommendations for the management of rheumatoid arthritis with synthetic and biological disease-modifying antirheumatic drugs: 2022 update. Ann Rheum Dis. 2023;82(1):3–18.

5. Genovese MC, Kremer J, Zamani O, Ludivico C, Krogulec M, Xie L, et al. Baricitinib in patients with refractory rheumatoid arthritis. N Engl J Med. 2016;374(13):1243–1252.

6. Genovese MC, Fleischmann R, Combe B, Hall S, Rubbert-Roth A, Zhang Y, et al. Safety and efficacy of upadacitinib in patients with active rheumatoid arthritis refractory to biologic disease-modifying anti-rheumatic drugs (SELECT-BEYOND): a double-blind, randomised controlled phase 3 trial. Lancet. 2018;391(10139):2513–2524.

7. Genovese MC, Kalunian K, Gottenberg JE, Mozaffarian N, Bartok B, Matzkies F, et al. Effect of filgotinib vs placebo on clinical response in patients with moderate to severe rheumatoid arthritis refractory to disease-modifying antirheumatic drug therapy: the FINCH 2 randomized clinical trial. JAMA. 2019;322(4):315–325.

8. Dörner T, Tanaka Y, Petri MA, et al. Baricitinib-associated changes in global gene expression during a 24-week phase II clinical systemic lupus erythematosus trial implicates a mechanism of action through multiple immune-related pathways. Lupus Sci Med. 2020;7(1):e000424.

9. Wampler Muskardin TL, Vashisht P, Dorschner JM, et al. Increased pretreatment serum IFN-β/α ratio predicts non-response to tumour necrosis factor α inhibition in rheumatoid arthritis. Ann Rheum Dis. 2016;75(10):1757–1762.

10. Cooles FAH, Anderson AE, Lendrem DW, et al. Interferon-α-mediated therapeutic resistance in early rheumatoid arthritis implicates epigenetic reprogramming. Ann Rheum Dis. 2022;81(9):1214–1223.

11. Cooles FAH, Isaacs JD. The interferon gene signature as a clinically relevant biomarker in autoimmune rheumatic disease. Lancet Rheumatol. 2022;4(1):e61–e72.

12. Castañeda-Delgado JE, Bastián-Hernandez Y, Macias-Segura N, et al. Type I interferon gene response is increased in early and established rheumatoid arthritis and correlates with autoantibody production. Front Immunol. 2017;8:285.

13. Londe AC, Fernandez-Ruiz R, Julio K, Appenzeller S, Niewold TB. Type I interferons in autoimmunity: implications in clinical phenotypes and treatment response. J Rheumatol. 2023;50(9):1103–1113.

14. Stockfelt M, Castegren M, Joutsi-Korhonen L, et al. Plasma interferon-α is associated with double-positivity for autoantibodies but is not a predictor of remission in early rheumatoid arthritis: a spin-off study of the NORD-STAR randomized clinical trial. Arthritis Res Ther. 2021;23(1):287.

