## Supplementary Material (Methods, Tables S1-S8, Figures S1-S5) for "Baseline ESR–CRP difference (D score) and JAK inhibitor discontinuation for loss of efficacy after biologic failure in rheumatoid arthritis"

### Supplementary Methods

#### Study design and data

This was a single-centre retrospective cohort study of 24 consecutive patients with rheumatoid arthritis who initiated a Janus kinase inhibitor (JAKi) after inadequate response to biological DMARDs (bDMARDs). The data cutoff date was 20 November 2025.

#### Outcome definitions and censoring

The primary outcome was discontinuation due to loss of efficacy (LOE), defined as discontinuation explicitly attributed to insufficient clinical response in the treating physician's record. Discontinuations for reasons other than LOE (e.g., adverse events) were treated as censored at the discontinuation date. Ongoing treatments were censored at the data cutoff date. Because adverse event discontinuation can act as a competing risk, we additionally performed an all-cause discontinuation analysis (LOE or adverse event discontinuation) as a sensitivity analysis.

#### Biomarker definition (D score)

Baseline ESR (mm/h) and CRP were obtained on the JAKi initiation date or within 14 days prior, using the closest available measurement. CRP values recorded in mg/dL were converted to mg/L by multiplying by 10. The D score was defined as ESR (mm/h) minus CRP (mg/L). Patients were dichotomised using the cohort median D score (20.3): High D (≥20.3) and Low D (<20.3).

#### Statistical analyses

Time-to-event analyses used Kaplan–Meier curves and log-rank tests. Cox proportional hazards models were fit using the Efron method for ties. Hazard ratios (HRs) are presented with 95% confidence intervals (CI). Proportional hazards assumptions were assessed using Schoenfeld residual plots; the corresponding p value in Supplementary Figure S2 is based on an approximate correlation test and should be interpreted cautiously given the small number of events.

#### Additional and sensitivity analyses

Sensitivity analyses included (i) all-cause discontinuation (LOE or adverse events) as the event (Supplementary Figure S3), (ii) adjustment for haemoglobin to address potential confounding of ESR by anaemia (Supplementary Table S5), (iii) an exploratory ESR×CRP quadrant analysis based on cohort medians to visualise concordant and discordant patterns (Supplementary Table S6 and Figure S4), and (iv) adjustment for, and exclusion of, patients treated with a preceding IL-6 receptor inhibitor to address potential CRP suppression (Supplementary Table S8).

#### Discrimination metrics

As exploratory analyses, discrimination was evaluated using Harrell's C-index and a pragmatic fixed-time 1-year AUC. For the 1-year AUC, patients censored before day 365 were excluded; the outcome was defined as LOE by day 365. Risk-score directionality was standardised such that higher scores indicate higher risk (ESR and D score were multiplied by −1). Confidence intervals were derived using non-parametric bootstrap resampling (1000 iterations).

#### Software

Analyses were performed in Python using pandas, NumPy, SciPy, statsmodels, scikit-learn, Matplotlib, and python-docx.

### Supplementary Table Legends

**Supplementary Table S1.** Baseline characteristics stratified by D score group (high $D \geq 20.3$ vs low $D < 20.3$).

**Supplementary Table S2.** Univariable Cox proportional hazards models for discontinuation due to loss of efficacy (LOE).

**Supplementary Table S3.** Cox proportional hazards model adjusted for age.

**Supplementary Table S4.** Exploratory discrimination metrics (Harrell's C-index and 1-year AUC) with bootstrap 95% confidence intervals.

**Supplementary Table S5.** Cox model including D score and haemoglobin.

**Supplementary Table S6.** ESR×CRP quadrant analysis (median cutoffs), showing the distribution of LOE and adverse-event discontinuations.

**Supplementary Table S7.** Preceding biologic DMARD classes before JAK inhibitor initiation.

**Supplementary Table S8.** Sensitivity analyses considering preceding IL-6 receptor inhibitor exposure.

### Supplementary Figure Legends

**Supplementary Figure S1.** Discrimination bar plots for ESR, CRP, and D score: Harrell's C-index and 1-year AUC.

**Supplementary Figure S2.** Schoenfeld residual plot for D score in the univariable Cox model (approximate proportional hazards test p value shown).

**Supplementary Figure S3.** Kaplan–Meier curves for all-cause discontinuation (LOE or adverse events), stratified by D score group.

**Supplementary Figure S4.** Kaplan–Meier curves for LOE-related discontinuation across ESR×CRP quadrant groups using median cutoffs.

**Supplementary Figure S5.** Scatter plot of baseline ESR versus CRP (mg/L) with median cutoffs, indicating LOE events, adverse-event discontinuation, and censoring.

**Supplementary Table S1. Baseline characteristics stratified by D score group.**

| Characteristic | High D (n=12) | Low D (n=12) | P value |
|---|---|---|---|
| Age, years | 72.5 (64.0 to 75.5) | 67.5 (54.2 to 73.2) | 0.173 |
| RA duration, years | 6.1 (1.9 to 16.4) | 3.6 (2.3 to 6.7) | 0.470 |
| ESR, mm/h | 73.5 (43.0 to 87.2) | 8.0 (2.0 to 18.2) | <0.001 |
| CRP, mg/L | 19.4 (4.9 to 35.6) | 0.7 (0.2 to 3.4) | 0.035 |
| D score (ESR - CRP) | 43.1 (30.9 to 61.9) | 2.9 (1.6 to 9.9) | <0.001 |
| Haemoglobin, g/dL | 12.7 (11.7 to 13.0) | 12.8 (12.2 to 13.3) | 0.603 |
| Prednisolone, mg/day | 4.0 (0.0 to 8.1) | 1.0 (0.0 to 5.4) | 0.378 |
| Prior bDMARD count | 3.0 (1.0 to 3.0) | 2.0 (1.0 to 3.0) | 0.322 |
| Female sex | 10/12 (83.3%) | 11/12 (91.7%) | 1.000 |
| RF positive | 11/12 (91.7%) | 9/12 (75.0%) | 0.590 |
| ACPA positive | 11/12 (91.7%) | 7/12 (58.3%) | 0.155 |
| Concomitant MTX | 6/12 (50.0%) | 5/12 (41.7%) | 1.000 |

**Supplementary Table S2. Univariable Cox models for LOE-related discontinuation.**

| Variable | Unit | HR | 95% CI | P value |
|---|---|---|---|---|
| D score | per 10 units | 0.47 | 0.29 to 0.76 | 0.002 |
| ESR | per 10 mm/h | 0.73 | 0.52 to 1.02 | 0.066 |
| CRP | per 10 mg/L | 1.00 | 0.73 to 1.36 | 0.984 |
| Age | per 1 year | 0.97 | 0.91 to 1.04 | 0.447 |

**Supplementary Table S3. Age-adjusted Cox model.**

| Variable | Unit | HR | 95% CI | P value |
|---|---|---|---|---|
| D score | per 10 units | 0.41 | 0.22 to 0.74 | 0.003 |
| Age | per 1 year | 0.93 | 0.85 to 1.03 | 0.156 |

**Supplementary Table S4. Exploratory discrimination metrics (C-index and 1-year AUC).**

| Biomarker | C-index | C-index 95% CI (lo) | C-index 95% CI (hi) | 1-year AUC | AUC 95% CI (lo) | AUC 95% CI (hi) |
|---|---|---|---|---|---|---|
| D score | 0.89 | 0.70 | 1.00 | 0.92 | 0.73 | 1.00 |
| ESR | 0.73 | 0.49 | 0.95 | 0.78 | 0.49 | 1.00 |
| CRP | 0.49 | 0.21 | 0.76 | 0.43 | 0.12 | 0.75 |

**Supplementary Table S4 (continued).**

| AUC_n_used | AUC_events_by_1y |
|---|---|
| 18 | 6 |
| 18 | 6 |
| 18 | 6 |

**Supplementary Table S5. Cox model including D score and haemoglobin.**

| Variable | Unit | HR | 95% CI | P value |
|---|---|---|---|---|
| D score | per 10 units | 0.53 | 0.33 to 0.87 | 0.012 |
| Haemoglobin | per 1 g/dL | 0.77 | 0.49 to 1.20 | 0.248 |

**Supplementary Table S6. ESR×CRP quadrant analysis (median cutoffs).**

| Quadrant | n (LOE events) | AE events |
|---|---|---|
| High ESR / High CRP | 10 (2) | 0 |
| High ESR / Low CRP | 2 (0) | 1 |
| Low ESR / High CRP | 2 (0) | 0 |
| Low ESR / Low CRP | 10 (5) | 1 |

**Supplementary Table S7. Preceding bDMARD classes.**

| Class | Drug | n (%) |
|---|---|---|
| CTLA4-Ig | ABT | 8 (33.3%) |
| IL-6 receptor inhibitor | SAR | 2 (8.3%) |
| IL-6 receptor inhibitor | TCZ | 2 (8.3%) |
| TNF inhibitor | OZR | 4 (16.7%) |
| TNF inhibitor | ADA | 2 (8.3%) |
| TNF inhibitor | ADA-BS | 2 (8.3%) |
| TNF inhibitor | GLM | 2 (8.3%) |
| TNF inhibitor | CZP | 1 (4.2%) |
| TNF inhibitor | ETN | 1 (4.2%) |

**Supplementary Table S8. Sensitivity analyses for preceding IL-6R inhibitor exposure.**

| Model | Variable | Unit | HR | 95% CI | P value |
|---|---|---|---|---|---|
| Adjusted for preceding IL-6R inhibitor | D score | per 10 units | 0.47 | 0.29 to 0.76 | 0.002 |
| Adjusted for preceding IL-6R inhibitor | Preceding IL-6R inhibitor | yes vs no | 1.05 | 0.17 to 6.39 | 0.957 |
| Excluded preceding IL-6R inhibitor | D score | per 10 units | 0.51 | 0.32 to 0.83 | 0.006 |

**Supplementary Figure S1. Discrimination bar plots for ESR, CRP, and D score.**

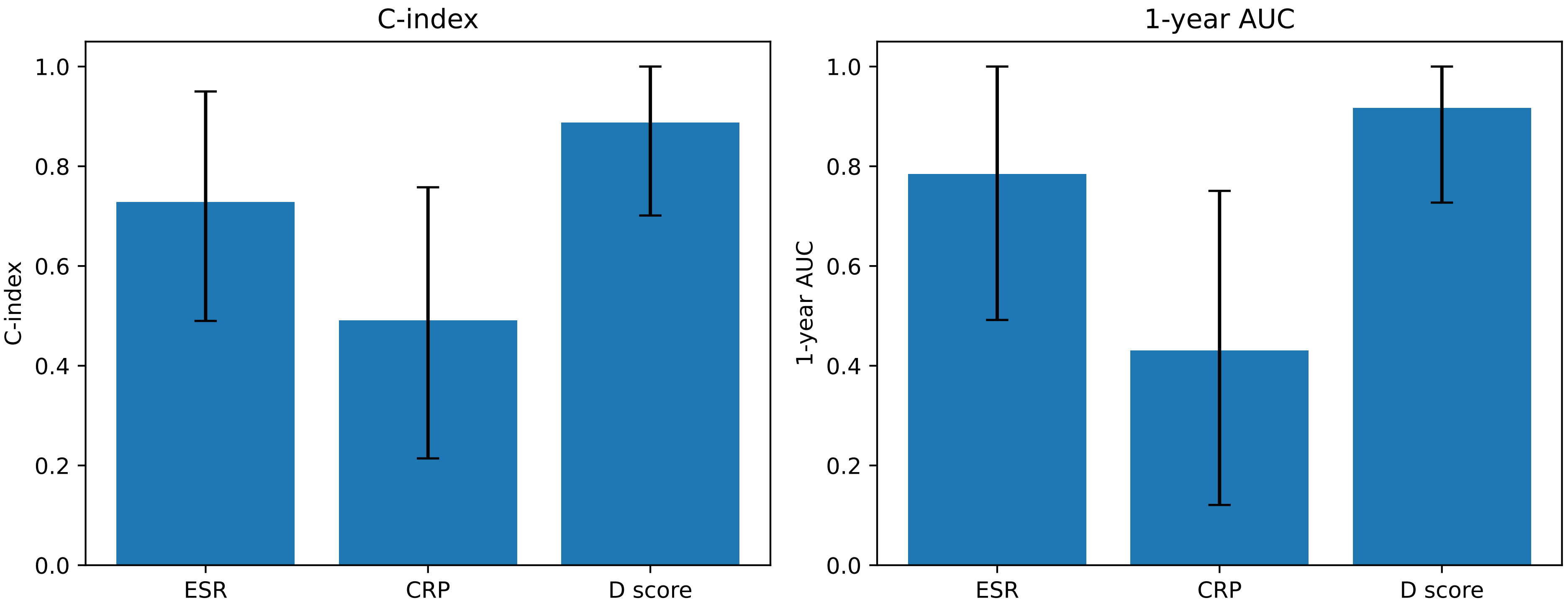

**Supplementary Figure S2. Schoenfeld residual plot for D score.**

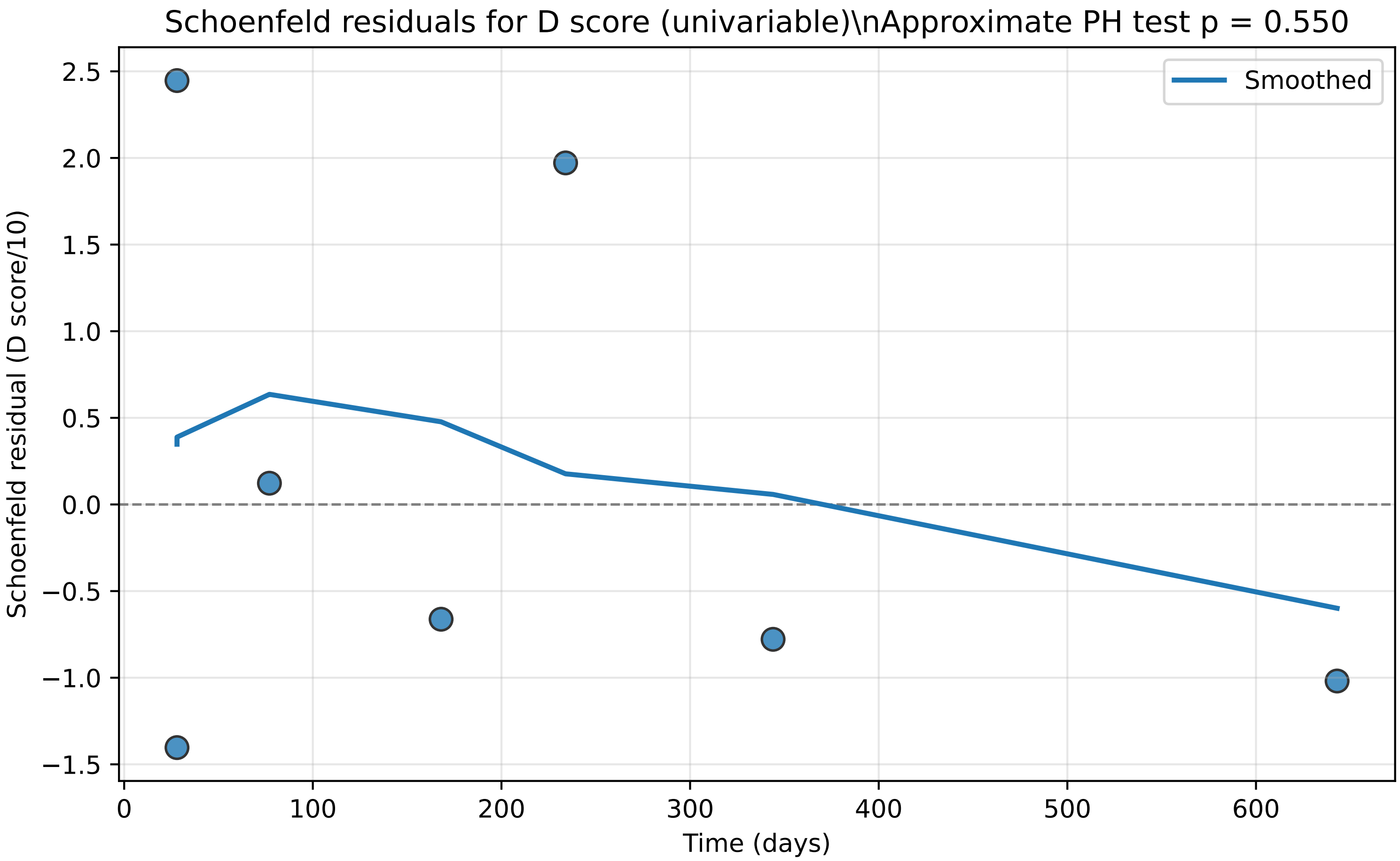

**Supplementary Figure S3. Kaplan–Meier curves for all-cause discontinuation.**

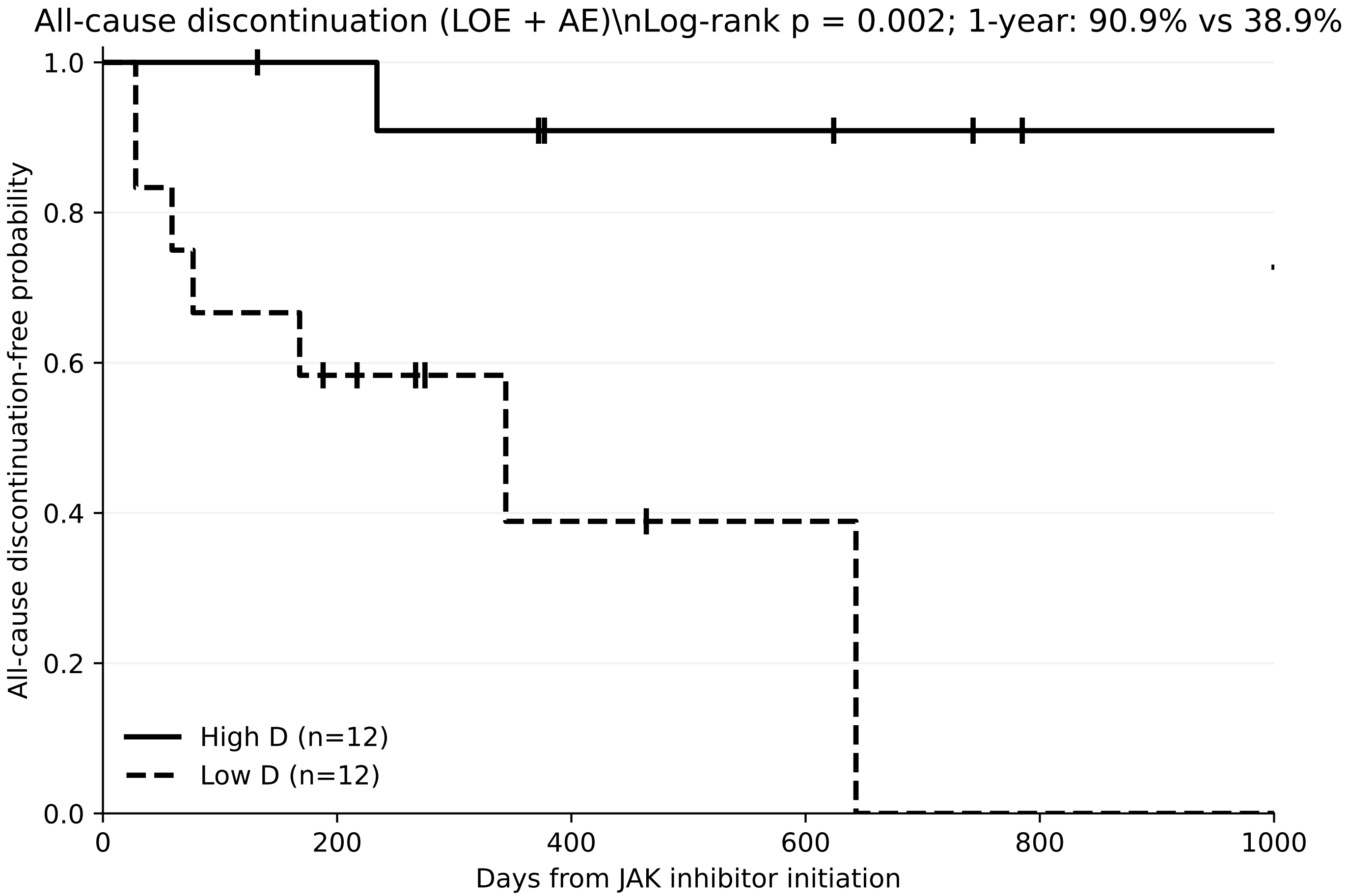

**Supplementary Figure S4. Kaplan–Meier curves by ESR×CRP quadrant groups.**

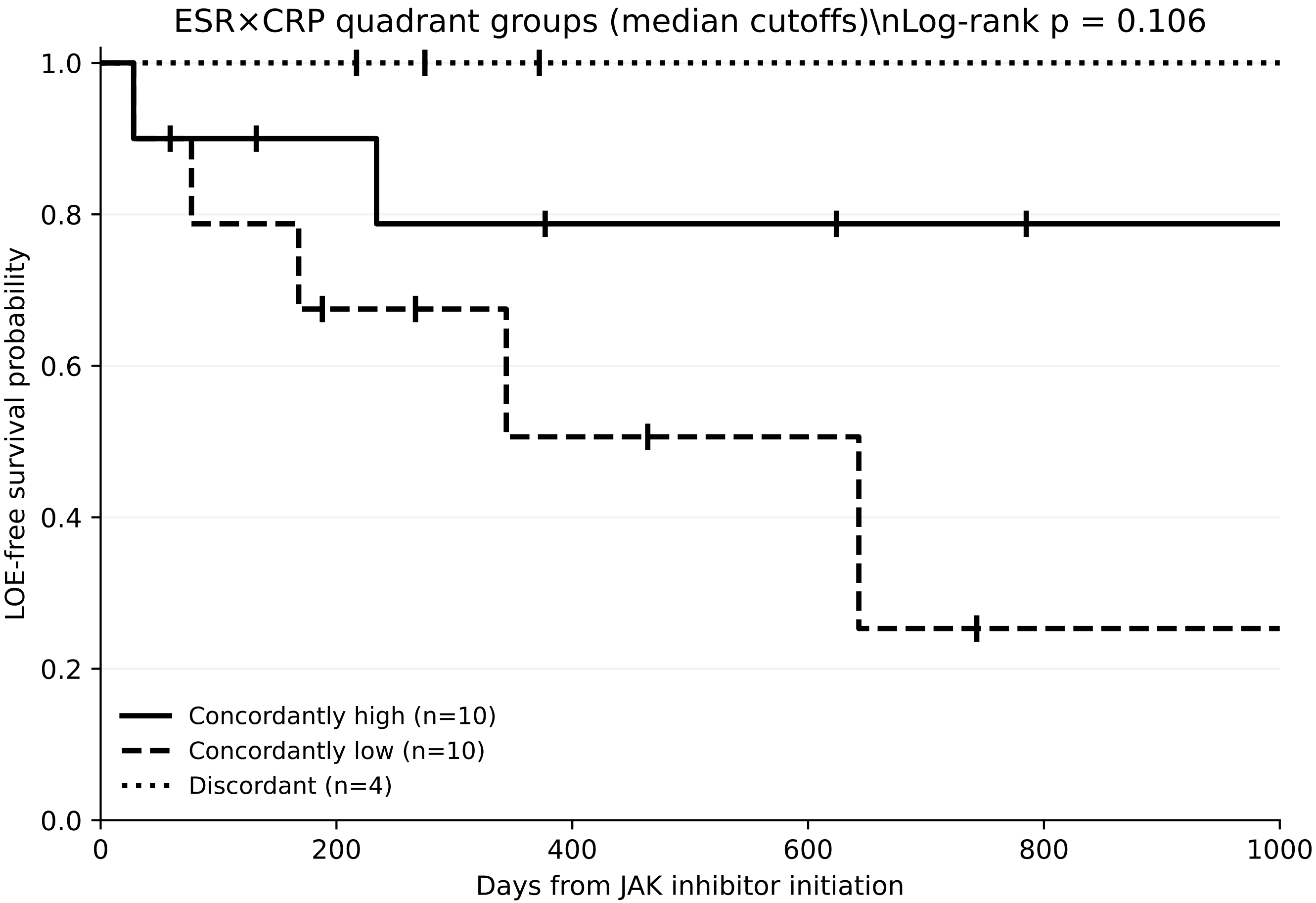

**Supplementary Figure S5. Scatter plot of baseline ESR vs CRP (mg/L).**

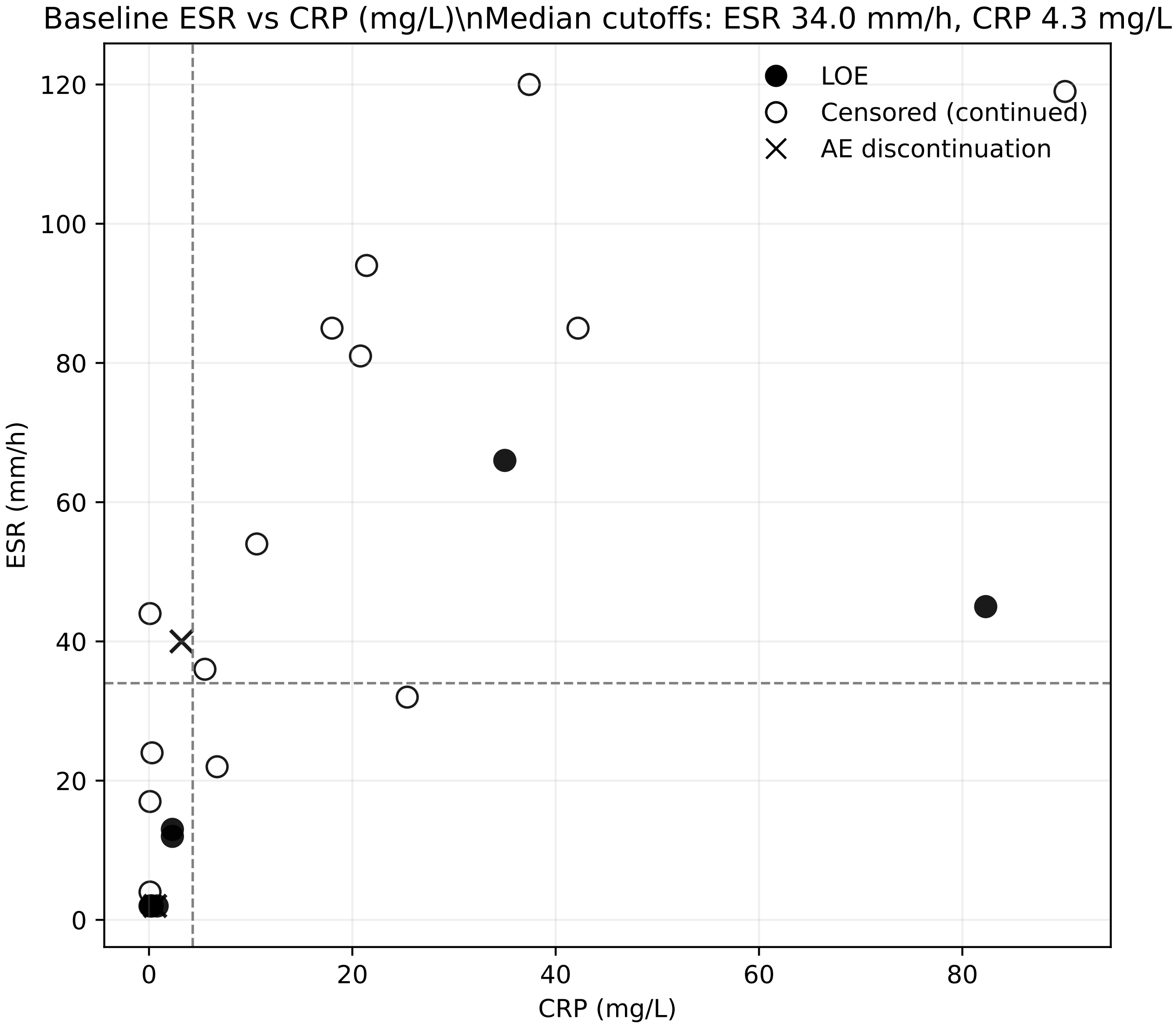
